# Estimating the Impact of Non-Pharmaceutical Interventions on COVID Mortality Using Reductions in Influenza Mortality as an Effect Indicator

**DOI:** 10.1101/2024.03.06.24303834

**Authors:** Robert Morris

## Abstract

This study uses influenza mortality reduction (IMR) as an indicator of the aggregate effect of non-pharmaceutical interventions (NPI’s) on the spread of respiratory infections to assess their impact on COVID mortality.

Age-adjusted COVID mortality for US states were modeled using four variables: COVID mortality prior to introduction of NPI’s, vaccination rates, IMR relative to 2016-2019, and population density.

A simple linear model of the entire pandemic with only these variables explained 65% of the variability in COVID mortality with IMRs affecting the first two years and vaccination having an impact in the second and third year. A counterfactual model of no NPI’s suggests they prevented 850,000 COVID related deaths in the United States.

These results support the use of IMR’s as an indicator of the aggregate impact of NPIs for controlling transmission of respiratory infections, including COVID and suggest that COVID mortality would have been almost 75% higher without them.

## Background

Since the outset of the COVID pandemic, non-pharmaceutical interventions (NPIs) to protect public health have come under heavy criticism for their impact on everything from the economy^1^ to mental health^2–4^ to education^5,6^. Furthermore, almost every intervention has, at some point, been declared ineffective, including masking,^7,8^ routine testing,^9^ school closures,^10^ and business “lockdowns”.^11,12^

Meta-analyses of studies of the efficacy of individual NPIs have tended to find beneficial effects^13–15^ with a few prominent exceptions.^7,8^ Closures of businesses and schools, limits on social gathering, travel restrictions, social distancing rules, masking mandates, and other NPI’s act in concert, often synergistically, to reduce the transmission of respiratory infections. Some protect the individual from exposure in the community, some protect the community from the infected individual, and some do both. Also, the effectiveness of NPI’s depends on compliance, which is difficult to quantify. How, then, do we evaluate the combined impact of these interventions on the transmission of COVID?

In an ideal natural experiment, we would have two isolated regions experiencing epidemic conditions that are identical in every way except for fully quantified and controlled differences in NPIs. Alternatively, we might have historical data for a particular disease and could examine changes in incidence and mortality after interventions were imposed. No such natural experiment occurred and, because COVID is a new human disease, we have no historical data. All of this makes the aggregate impact of NPI’s on COVID difficult to assess directly.

However, the impact of these NPIs was not limited to COVID. Interventions designed to stop one respiratory pathogen will stop others as well. Therefore, the extent to which these NPIs halted the spread of respiratory pathogens with similar patterns of airborne transmission may provide a surrogate for the efficacy of NPIs for COVID. By far the best characterized of these diseases is influenza.

The marked seasonal patterns of influenza incidence and mortality have been measured for decades. As a result, the expected annual influenza mortality and the variability in that mortality are well established. Multiple studies have noted the dramatic, global decline in influenza incidence and mortality^16–18^ during the COVID pandemic and suggested an association with NPIs. In the United States, influenza mortality rates for the first two complete flu seasons (2020-22) were 80% below historic rates,^19^ as can be seen in Figure 1. The sharpness of this decline and the fact that flu mortality rose back to pre-pandemic levels once precautions were lifted in 2022-23 makes it unlikely that the drop was due to any change in the transmissibility or infection fatality rate of the virus. Influenza vaccination rates did rise 7% above historical averages during the pandemic^20^ (probably due to concurrent vaccination with COVID), but this cannot explain an 80% drop in influenza mortality.

**Figure 1.**
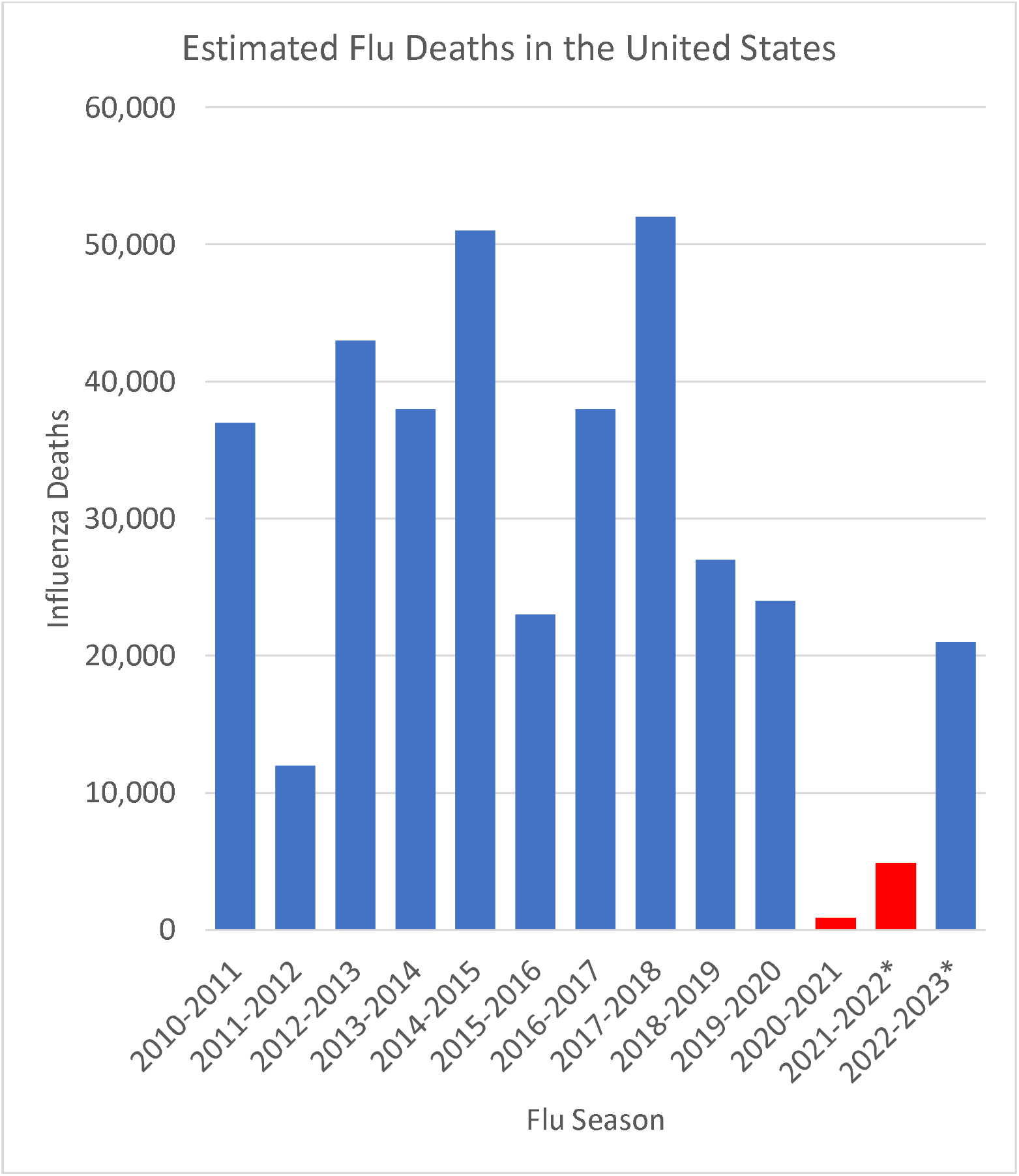
CDC estimates of deaths from influenza for past 13 flu seasons. Note that the CDC did not provide an estimate for the 2020-2021season because the mortality rates were too low for their estimation procedures, which seek to account for unreported cases, so the number provided is the actual count.

It appears that something changed during the pandemic that resulted in a dramatic drop in respiratory disease transmission. By far the most likely explanation of this is COVID NPI’s. That suggests that the extent to which influenza mortality decreased from expected levels represents a drop in respiratory disease transmission and may provide an indicator of the effectiveness of COVID NPI’s. The current study explores the association between the influenza mortality reduction (IMR) and COVID mortality for US states.

## Methods

The incidence and mortality rates of COVID (or any infectious disease) in a state are a function of four things: 1.) the rate of introduction of new cases to the state, 2.) the probability of infected individuals encountering uninfected community members under normal circumstances, 3.) non-pharmaceutical interventions (NPIs) taken to reduce the likelihood of transmissions, either by limiting social interactions (school and business closures, social distancing) or by reducing risk associated with those interactions (masks, barriers, hand washing), and 4.) immunity through vaccination or prior infection.

Four variables were used to represent these four factors:

1. **Initial COVID mortality:** State specific, weekly COVID mortality data were obtained from the CDC COVID Data Tracker and used to determine initial COVID mortality rates.^21^ The first state shutdowns were implemented on March 15 of 2020^22^. To account for the lag between implementation of NPIs and their impact on mortality, the COVID mortality rates for through the end of March, 2020 were used as an indicator of how severely a state was impacted by COVID prior to implementation of NPIs.
2. **Population density:** State population densities for 2020 were obtained from the United States Census Bureau^23^ to serve as an indicator of the baseline probability of social encounters within a state.
3. **IMR** for each state during the first two full flu seasons of the pandemic served as a surrogate for the impact of NPIs. To estimate IMR, weekly counts of influenza deaths for the period from 2016 through 2023 were abstracted from the CDC FluView System^24^ for each state. Influenza mortality rates for each state were calculated for pre-COVID flu seasons from 2016 through 2019 and for the two flu seasons during the pandemic, 2020-21 and 2021-22. Note that the 2019-2020 flu season was not considered because it included periods before and after implementation of NPIs and it was ending naturally, just as COVID was beginning. IMR was calculated as

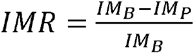

where: IM_B_ = the average influenza mortality rate for the three seasons prior to the pandemic, and IM_P_ = the average influenza mortality for flu seasons during the pandemic. Values of IMR were calculated for the 2020-2021 flu season, for the 2021-2022 flu season, and for the combined flu seasons.
4. **Vaccination Rate** (per cent of population fully vaccinated for COVID) provided a measure of population immunity. Vaccination rates for each state were obtained from the CDC with vaccination defined as receipt of two initial doses.^21^ Rates were determined at three time points: September of 2021, September of 2022, and September of 2023.

The outcome variable for this analysis was state-specific COVID mortality rates. Mortality was used rather than incidence because it is far less susceptible to variations in case ascertainment. COVID mortality rates were determined based on age specific COVID mortality rates from CDC for four time periods based on the flu seasons to match with the IMR data. The time periods were Flu Season 1 (January-September 2020), Flu Season 2 (October 2020 through September 2021), Flu Season 3 (October 2021 through September 2022), and the full pandemic (January 2020 through September 2023).^25^ State age distributions for 2020 were obtained from the US Census Bureau^26^ and COVID mortality was adjusted to the 2020 US national population.

A multiple linear regression model was used to estimate state specific, age adjusted COVID mortality as a function of population density, initial COVID mortality, IMR, and vaccination rates for each time period. Note that model for Season 1 did not include vaccination, since the vaccine was not introduced until October of 2020. It also used the IMR for the 2020-21 flu season as a surrogate for 2020 NPIs, because, as noted above, the 2019-2020 flu season was largely over when the pandemic began. The assumption is that state practices for 2020-2021 were an extension of practices introduced in 2020. All statistical analyses described above were conducted using the STATA statistical package. Standardized regression coefficients were calculated to compare the relative impact of the independent variables. Variance inflation factors were calculated to assess the impact of collinearity on the models.

The resulting models were used to evaluate the counterfactual cases of non-intervention by setting the relevant variable to zero for each state, determining predicted deaths for each state, and summing the results. Because there is no way to directly calculate the confidence interval for the sum of the estimates, Monte Carlo simulations with 10,000 repetitions were used to estimate the distribution of the aggregated mortality predictions and extract confidence intervals. The Monte Carlo models were written in Python (code available on request). The two counterfactual cases evaluated were no NPIs (IMR=0) and no vaccination or NPIs. To assess the counterfactual case of strong public health interventions, a simulation was run based on all states matching the IMR and vaccination rates of the top performing states.

## Results

As listed in Table 1, state-level influenza mortality rates were an average of 78% lower during the two full flu seasons of the pandemic, 2020-21 and 2021-22, as compared to the three full flu seasons prior to the pandemic, 2016-17, 2017-18, and 2018-19. The IMR ranged from 49% (North Dakota) to 94% (Washington).

**Table 1.** Vaccination, population density, influenza and COVID mortality rates (deaths/100,000).

| State | Pre-Covid<br>Influenza<br>Mortality<br>Rate | 2020-2022<br>Influenza<br>Mortality<br>Rate | Total<br>Infection<br>Mortality<br>Reduction<br>(IMR) | Vaccination<br>Rate (2023) | Population<br>Density<br>(Per mi<sup>2</sup>) | Initial<br>Covid<br>Mortality | Age-Adjusted<br>Covid<br>Mortality |
| --- | --- | --- | --- | --- | --- | --- | --- |
| Alabama | 2.55 | 0.77 | 71.0% | 53% | 99 | 0.52 | 422 |
| Alaska | 2.73 | 0.95 | 60.6% | 65% | 1 | 3.59 | 269 |
| Arizona | 2.35 | 0.51 | 77.8% | 66% | 63 | 0.35 | 408 |
| Arkansas | 4.09 | 0.88 | 77.7% | 57% | 58 | 0.86 | 418 |
| California | 2.51 | 0.28 | 88.8% | 75% | 254 | 0.07 | 296 |
| Colorado | 3.15 | 0.80 | 73.4% | 74% | 56 | 0.45 | 302 |
| Connecticut | 3.59 | 0.40 | 88.2% | 83% | 745 | 0.73 | 310 |
| Delaware | 2.97 | 0.34 | 87.7% | 74% | 508 | 2.55 | 316 |
| Florida | 1.88 | 0.67 | 66.4% | 70% | 402 | 0.12 | 318 |
| Georgia | 1.58 | 0.57 | 67.4% | 58% | 186 | 0.24 | 387 |
| Hawaii | 3.07 | 0.35 | 87.6% | 82% | 227 | 1.84 | 117 |
| Idaho | 4.34 | 0.87 | 77.2% | 57% | 22 | 1.34 | 321 |
| Illinois | 2.80 | 0.35 | 86.3% | 72% | 231 | 0.21 | 305 |
| Indiana | 3.76 | 0.69 | 80.3% | 58% | 189 | 0.38 | 406 |
| Iowa | 5.38 | 1.00 | 79.8% | 65% | 57 | 0.82 | 317 |
| Kansas | 5.25 | 1.07 | 79.0% | 66% | 36 | 0.90 | 346 |
| Kentucky | 4.52 | 0.80 | 81.5% | 60% | 114 | 0.58 | 457 |
| Louisiana | 2.52 | 0.60 | 75.7% | 55% | 108 | 0.58 | 397 |
| Maine | 5.16 | 0.68 | 85.6% | 84% | 44 | 1.89 | 190 |
| Maryland | 2.03 | 0.37 | 83.0% | 80% | 636 | 0.43 | 305 |
| Massachusetts | 3.27 | 0.79 | 75.3% | 85% | 901 | 0.38 | 281 |
| Michigan | 3.18 | 0.65 | 79.3% | 63% | 178 | 0.26 | 349 |
| Minnesota | 3.68 | 0.61 | 82.4% | 72% | 72 | 0.46 | 266 |
| Mississippi | 2.77 | 1.17 | 57.3% | 54% | 63 | 0.90 | 508 |
| Missouri | 4.55 | 0.78 | 81.5% | 59% | 90 | 0.43 | 370 |
| Montana | 4.86 | 1.06 | 76.7% | 59% | 8 | 2.33 | 332 |
| Nebraska | 4.77 | 1.16 | 72.9% | 67% | 26 | 1.33 | 300 |
| Nevada | 1.62 | 0.53 | 69.1% | 64% | 28 | 0.82 | 423 |
| New Hampshire | 4.04 | 0.82 | 77.1% | 72% | 154 | 1.88 | 202 |
| New jersey | 1.94 | 0.25 | 86.9% | 79% | 1263 | 0.28 | 377 |
| New Mexico | 2.79 | 0.76 | 74.7% | 76% | 18 | 1.25 | 413 |
| New York | 1.64 | 0.35 | 78.2% | 81% | 429 | 0.13 | 385 |
| North Carolina | 3.47 | 0.44 | 86.7% | 67% | 215 | 0.24 | 332 |
| North Dakota | 3.32 | 1.66 | 49.0% | 59% | 11 | 3.36 | 404 |
| Ohio | 3.46 | 0.49 | 85.1% | 61% | 289 | 0.22 | 398 |
| Oklahoma | 4.51 | 1.57 | 64.1% | 61% | 58 | 0.65 | 487 |
| Oregon | 5.55 | 0.34 | 92.8% | 73% | 44 | 0.62 | 206 |
| Pennsylvania | 3.44 | 0.73 | 77.5% | 74% | 291 | 0.20 | 356 |
| Rhode Island | 4.78 | 0.50 | 88.4% | 88% | 1061 | 2.40 | 328 |
| South Carolina | 3.54 | 0.61 | 82.1% | 60% | 170 | 0.49 | 387 |
| South Dakota | 5.29 | 1.69 | 64.6% | 67% | 12 | 2.87 | 369 |
| Tennessee | 3.31 | 1.16 | 65.8% | 56% | 168 | 0.37 | 446 |
| Texas | 2.15 | 0.67 | 68.7% | 64% | 112 | 0.09 | 433 |
| Utah | 2.07 | 0.50 | 74.8% | 67% | 40 | 0.77 | 252 |
| Vermont | 5.41 | 0.46 | 90.7% | 86% | 70 | 4.07 | 129 |
| Virginia | 2.36 | 0.47 | 79.1% | 77% | 219 | 0.30 | 281 |
| Washington | 4.61 | 0.26 | 93.6% | 76% | 116 | 0.34 | 207 |
| West Virginia | 5.07 | 1.30 | 73.1% | 60% | 75 | 1.49 | 404 |
| Wisconsin | 3.91 | 0.52 | 85.8% | 68% | 109 | 0.45 | 271 |
| Wyoming | 3.48 | 0.77 | 75.0% | 53% | 6 | 4.51 | 326 |
| Average | 3.50 | 0.72 | 78% | 68% | 207 | 1.05 | 337 |

As shown in Table 2, COVID mortality had a strong negative correlation with IMR and vaccination rates. IMR is also strongly correlated with vaccination rates. Variance inflation factors were calculated for all of the models described below and the highest value observed was 1.97 suggesting collinearity should not be a problem for the regression models.^27^

**Table 2.** Pairwise Pearson correlation coefficients for model variables.

|  | pop<br>density | Initial | <u>COVID Mortality</u> |  |  |  |  | <u>IMR</u> |  | <u>Vaccination</u> |  |
| --- | --- | --- | --- | --- | --- | --- | --- | --- | --- | --- | --- |
|  |  |  | Season<br>1 | Season<br>2 | Season<br>3 | All | Season<br>2 | Season<br>3 | All | 2021 | 2022 |
| **COVID Mortality** |  |  |  |  |  |  |  |  |  |  |  |
| Initial* | -0.18 | 1.00 |  |  |  |  |  |  |  |  |  |
| Season 1 | 0.74 | -0.40 | 1.00 |  |  |  |  |  |  |  |  |
| Season 2 | -0.22 | -0.16 | 0.17 | 1.00 |  |  |  |  |  |  |  |
| Season 3 | -0.32 | 0.05 | -0.19 | 0.65 | 1.00 |  |  |  |  |  |  |
| All | -0.04 | -0.29 | 0.36 | 0.89 | 0.58 | 1.00 |  |  |  |  |  |
| **IMR** |  |  |  |  |  |  |  |  |  |  |  |
| Season 2 | 0.20 | 0.15 | -0.21 | -0.59 | -0.11 | -0.60 | 1.00 |  |  |  |  |
| Season 3 | 0.30 | -0.24 | 0.08 | -0.41 | -0.32 | -0.43 | 0.44 | 1.00 |  |  |  |
| All | 0.34 | -0.18 | 0.02 | -0.53 | -0.29 | -0.54 | 0.70 | 0.94 | 1.00 |  |  |
| **Vaccination** |  |  |  |  |  |  |  |  |  |  |  |
| 9/2021 | 0.54 | -0.02 | 0.18 | -0.70 | -0.52 | -0.62 | 0.46 | 0.46 | 0.55 | 1.00 |  |
| 9/2022 | 0.57 | 0.02 | 0.16 | -0.73 | -0.54 | -0.65 | 0.49 | 0.44 | 0.55 | 0.97 | 1.00 |
| 9/2023 | 0.57 | 0.03 | 0.17 | -0.72 | -0.53 | -0.64 | 0.50 | 0.43 | 0.54 | 0.97 | 0.99 |

\* The time periods are defined as follows: Initial includes only deaths through March 31,2020. Season 1 includes January through September 2020. Season 2 includes October 2020 through September 2021, Season 3 includes October 2021 through September 2022, All includes January 2020 through September 2023.

Multiple linear regression model results are provided in Table 3 for the four different time periods. In the period prior to the introduction of the vaccine the model provides an excellent fit (r^2^ = 0.70) with COVID mortality determined primarily by population density and the NPI surrogate, IMR, with SRCs of 0.77 and −0.33 respectively. IMR had a similarly strong association with COVID mortality in the second season (SRC = −0.31), while the effect of population density faded (SRC = 0.18). Vaccination rate had the strongest association of all variables in the model (SRC = −0.66). In the third season, the model r^2^ dropped to 0.24 and only vaccination rate had a significant impact on COVID mortality. When the full pandemic is considered, the model predicts pandemic COVID mortality at the state level with an r^2^ of 0.65. Vaccination rates and IMR both had strong negative associations with COVID death rates [standardized regression coefficients (SRC) of −0.68 and −0,36 respectively]. Initial COVID mortality had a negative SRC of −0.27 and population density had an SRC of 0.45.

**Table 3.** Multiple linear regression results for US state, age adjusted COVID mortality rates as a function of Influenza Mortality Reduction (IMR), vaccination rate (at least 2 doses), COVID mortality for four time periods: Season 1 includes January through September 2020. Season 2 includes October 2020 through September 2021, Season 3 includes October 2021 through September 2022, The combined analysis includes January 2020 through September 2023.

| Period | State Characteristics | Coeff. | 95% C.I. |  | P>\|t\| | Standardized Coeff. |
| --- | --- | --- | --- | --- | --- | --- |
| 2020<br>Season 1<br>r<sup>2</sup>= 0.70 | Initial COVID Mortality | -6.32 | -11.1 | -1.49 | 0.011 | -0.212 |
|  | Population Density | 0.0917 | 0.0723 | 0.111 | <0.001 | 0.769 |
|  | IMR 2020-21 | -130 | -195 | -65 | <0.001 | -0.325 |
|  | Constant | 155 | 97 | 212 | <0.001 |  |
| 2020–21<br>Season 2<br>r<sup>2</sup> = 0.59 | Initial COVID Mortality | -4.33 | -13.4 | 4.74 | 0.342 | -0.091 |
|  | Population Density | 0.0336 | -0.0089 | 0.0761 | 0.118 | 0.178 |
|  | IMR 2020-21 | -197 | -332 | -62.7 | 0.005 | -0.311 |
|  | Vaccination Rate | -414 | -569 | -260 | <0.001 | -0.655 |
|  | Constant | 551 | 439 | 663 | <0.001 |  |
| 2021-22<br>Season 3<br>r2= 0.24 | Initial COVID Mortality | 1.076 | -6.411 | 8.563 | 0.774 | 0.039 |
|  | Population Density | -0.0013 | -0.0363 | 0.0336 | 0.939 | -0.012 |
|  | IMR 2020-21 | -20.3 | -84.6 | 44.0 | 0.528 | -0.092 |
|  | Vaccination Rate | -162 | -273 | -51.1 | 0.005 | -0.490 |
|  | Constant | 226 | 160 | 291 | <0.001 |  |
| 2020-2023<br>Season 1-3<br>r2=0.65 | Initial COVID Mortality | -20.8 | -34.7 | -6.8 | 0.004 | -0.268 |
|  | Population Density | 0.1248 | 0.0589 | 0.191 | <0.001 | 0.404 |
|  | IMR 2020-22 | -336 | -525 | -147 | 0.001 | -0.370 |
|  | Vaccination Rate | -5.78 | -7.88 | -3.67 | <0.001 | -0.657 |
|  | Constant | 986 | 843 | 1129 | <0.001 |  |

**Table 4.**
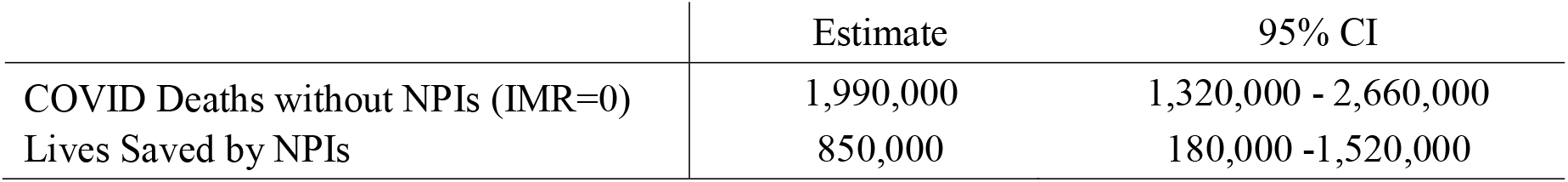
Model estimates of lives saved by NPI’s (based on IMR as an indicator). Mortality data in model based on values when US COVID mortality was 1.04 million.

|  | Estimate | 95% CI |
| --- | --- | --- |
| COVID Deaths without NPIs (IMR=0) | 1,990,000 | 1,320,000 - 2,660,000 |
| Lives Saved by NPIs | 850,000 | 180,000 -1,520,000 |

Figure 2 provides a graph of the actual age-adjusted COVID mortality as a function of the Modeled COVID mortality. The graph demonstrates the close fit of the model to the actual data and the absence of outliers.

**Figure 2.**
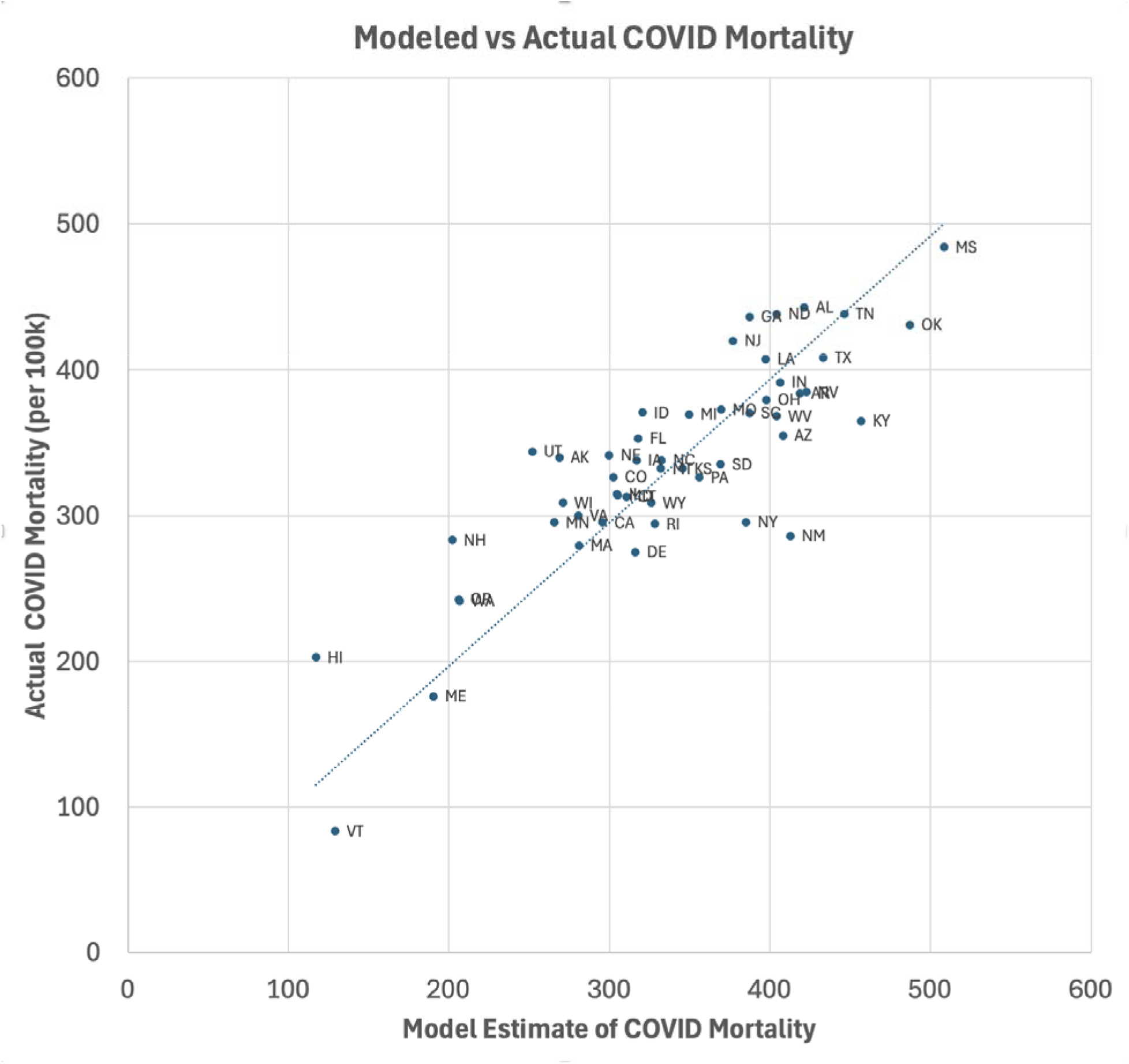
Age-adjusted COVID mortality as compared to model estimates for US states. Age adjusted COVID Mortality through the September of 2023 vs multiple linear regression model estimates using influenza mortality reduction [as an indicator of the aggregate effectiveness of non-pharmaceutical interventions (NPI’s)], COVID vaccination rates, population density, and initial COVID mortality (through March 2020). Model r^2^ =0.65.

The simulation of the counterfactual case with IMR set to zero estimated COVID mortality of 1.99 million (95% CI of (1.32 – 2.66 million) through September of 2023. The actual cumulative COVID mortality at that point was 1.14 million deaths, suggesting that NPIs saved 850,000 lives. If we set both IMR and vaccination to zero, the COVID mortality estimate rises to 3.39 million (95% CI of (2.93 – 3.86 million) suggesting that the combined preventative measures saved 2,350,000 lives.

On the other hand, if all states had matched the top IMR (94% for Washington), the model suggests we might have reduced deaths to 935,000, an 18% reduction in deaths from the observed number of 1.14 million. If, in addition, they had matched the vaccination rate for Rhode Island (88%), they would have reduced deaths to 535,000, a 53% reduction.

## Discussion

The current study demonstrates a strong association between IMR and reduced COVID mortality, both before and after the introduction of COVID vaccines. The most likely explanation of this association is that the NPIs introduced to control COVID also controlled influenza. If IMR reflects the effectiveness of NPIs, the model results suggest they played a key role in limiting the impact of the pandemic. Using the resulting model to estimate the hypothetical case of no NPIs suggests that they prevented 850,000 COVID deaths. Any assertion of causality, however, requires careful consideration of the limitations of the model and alternative explanations observed associations.

During the first nine months of 2020, a model based on just three variables, initial COVID mortality, population density, and IMR explained 70% of the variability in state COVID mortality rates. Population density had the strongest association with COVID mortality through September of 2020.

The shift in model results for the 2020-21 flu season is consistent with a major effect of vaccination on reducing COVID mortality. The persistent association with IMR suggests ongoing benefits of NPIs despite calls for discontinuing them.^28^

By 2022, the model breaks down with r^2^ dropping to 0.24 and only vaccination remaining as a significant predictor of COVID mortality. By this point, the dynamics of the pandemic had changed dramatically. Proactive use of NPIs had diminished and COVID cases arrived in waves as new variants appeared. NPIs were often introduced in response to increases in COVID, obscuring any ongoing effectiveness. Also, many of the most vulnerable members of the population had died from COVID and many others who were unvaccinated had been infected and developed immunity that is not represented in the model. By September of 2022, there had been almost 100 million confirmed cases of COVID in the US and the introduction of free home test kits in January of 2022^29^ means that there were many more who had tested positive but had not reported the infection. All this would make vaccination rates only a partial measure of COVID immunity.

One somewhat surprising finding is that initial COVID mortality had a persistent negative association with total COVID mortality. This may reflect the effect of early adverse experience with the pandemic on both state government willingness to impose NPIs and individual and local compliance with those NPIs. Alternatively, it is possible that the states hardest hit by the initial wave of COVID were those with high levels of international travel, such as the northeast and the west coast, which also tend to be more open to government intervention and willing to impose NPIs. Finally, densely populated states may have had superior medical care, enhancing survival rates.

## Strengths and Limitations

It follows logically that NPIs intended to control the spread of COVID would also control the spread of other respiratory viruses. This is evident in data from the Seattle Flu Study, an active surveillance program put in place prior to the pandemic, which demonstrated sharp decreases in the incidence of all respiratory infections including influenza, respiratory syncytial virus, and non-COVID corona viruses during the pandemic.^30,31^ Because influenza incidence and mortality is routinely tracked with well-established surveillance systems, it has the potential to serve as an indicator of the combined effectiveness of NPIs for the COVID pandemic and for future respiratory disease pandemics.

Disease ascertainment must be considered when using surveillance data. Could an overstressed healthcare system simply have been testing less for influenza or were tests less sensitive? To minimize any possible problems with ascertainment, this study focused on influenza mortality rather than incidence. Failure to diagnose a fatal case of the flu correctly, even during the pandemic, seems unlikely given the well-established surveillance system and diagnostic tools for influenza.

Some have gone so far as to suggest that COVID deaths were actually misdiagnosed influenza deaths.^32^ If flu deaths were being misdiagnosed as COVID, we would expect the reduction in influenza mortality to have a strong positive correlation with COVID mortality rates, not the strong negative correlation observed in these data.

The greatest limitation to the use of IMRs and as an indicator of NPI effectiveness for COVID is that the 2019-2020 flu season was fading just as the COVID pandemic was beginning. This meant there was no direct surrogate for NPI effectiveness for the critical first months of the pandemic, when the most stringent NPI’s were imposed. Even though the IMR for the subsequent flu season was a poor substitute, it had a strong association COVID mortality, seeming to justify its use. If the errors introduced by this surrogate are random, a better measure would be expected to increase the observed association.

The fact that this is an ecological study conducted over relatively large geographic areas might raise concerns about the ecological fallacy. The fact that respiratory disease transmission happens at the community level makes this less of a concern. If implementation and compliance vary across the state, we can’t be certain that COVID mortality reductions were higher in high compliance communities but it is difficult to see a plausible alternative explanation.

Other factors that could affect COVID mortality that might not be captured in these data include travel and the introduction of COVID out of state visitors, IMR driven by reduced influenza incidence in surrounding states, and differences in the effectiveness in quality of medical care among states. For the most part, these factors are likely to result in random misclassification and decreases in observed associations.

The simulation model is speculative and is predicated on the assumption that observed effects are causal and associations can be projected for the extreme cases. The excellent fit of this model supports the legitimacy of its use to evaluate counterfactual situations. If influenza mortality reduction corresponds to the effectiveness of NPIs, setting IMR to zero would correspond to no use of NPIs.

Although this study supports the use of IMR as a surrogate measure of the combined effect of NPIs, it does not tell us the relative contribution of specific interventions, such as masks, lockdowns, or school closures. Studies of individual interventions are deceptively difficult to conduct properly because many of them operate at a community level, meaning the unit of randomization for any sort of intervention trial would need to be the community. Observational studies on the other hand tend to be complicated by the fact that multiple interventions are often used together. Nonetheless, further research to understand the effectiveness of specific NPI’s and combinations of NPI’s will be critical to refining management strategies for future epidemics of respiratory infectious disease.

## Conclusions

When the entire pandemic is considered, this relatively simple model provides a remarkably good fit to the data given the fact that the model relied on data aggregated over long time periods and large geographic areas. The resulting misclassification is most likely to obscure detectable effects. All four variables in the model are highly significant and an adjusted r^2^ of 0.65 with no outliers strongly supports the validity of IMR as a surrogate for the cumulative effect of NPIs. IMRs appear to have been effective through the first two years of the pandemic.

Counterfactual simulations suggested that, in the absence of NPIs, COVID mortality rates would have been ~70% percent higher than observed. Additional simulations suggest that, if all states had matched the IMR of the top performing state we might have reduced observed COVID mortality rates by ~20%.

Further research on IMRs as a surrogate for the aggregate effectiveness of NPIs is warranted as are studies to investigate the contribution of individual interventions to this cumulative effect.

## Data Availability

All data produced in the present study are available upon reasonable request to the author.

https://www.cdc.gov/flu/weekly/index.htm

https://covid.cdc.gov/covid-data-tracker

https://data.cdc.gov/NCHS/Provisional-COVID-19-Deaths-by-Sex-and-Age/9bhg-hcku/about_data

https://www2.census.gov/programs-surveys/decennial/2020/data/apportionment/population-density-data-table.pdf

## Key Messages for Social Media

1. Influenza mortality for US states dropped dramatically during the 2020-21 and 2021-22 flu seasons as compared to the 2016-2019 seasons. Greater drops in mortality were strongly correlated with lower COVID mortality.
2. The only plausible explanation for this influenza mortality reduction (IMR) is reduced transmission resulting from non-pharmaceutical interventions (NPIs) to prevent the spread of COVID.
3. A simple linear model including only IMR, vaccination rates, population density, and early COVID mortality explained 65% of state to state variability in COVID mortality and suggested that NPIs prevented 850,000 COVID related deaths in the US.

## About the Author

Robert Morris, MD, PhD, is an Affiliate Associate Professor at the University of Washington School of Public Health, Seattle, WA, USA

